# Factors associated with decision making on COVID-19 vaccine acceptance among college students in South Carolina

**DOI:** 10.1101/2020.12.03.20243543

**Authors:** Cheuk Chi Tam, Shan Qiao, Xiaoming Li

## Abstract

**Background:** COVID-19 vaccination could be a promising approach in controlling the pandemic, but its success strongly relies on the acceptance of vaccines among various populations including young adults who are especially vulnerable to COVID-19 due to their active lifestyle and perception of invulnerability. Vaccine acceptance decisions can be influenced by multiple factors and people may weigh these factors differently in their decision making. The current study aims to explore COVID-19 vaccine acceptance among college students in South Carolina and examine how they weigh these factors according to their levels of COVID-19 vaccine acceptance (i.e., acceptance, hesitance, refusal).

**Methods:** Online survey data were collected from 1062 college students in South Carolina between September and October 2020. Multivariate analysis of covariance was used to compare perceived importance of 12 factors affecting levels of vaccine acceptance, controlling for key demographic variables.

**Results:** About 60.6% of the college students reported they would definitely or likely take COVID-19 vaccine when available. Duration of vaccine protection, vaccine accessibility, and authoritative advice (e.g., if vaccination is recommended by school, government, or doctors) were considered important among the acceptance group; Negative consequences of vaccination and vaccine characteristics (i.e., ways the vaccine will be administered, and where the vaccine is made) were considered important by the refusal group; The hesitance group considered the same factors important as the refusal group did but also considered duration of vaccine protection and recommendation by school or doctors important.

**Conclusion:** Our findings suggest relatively low vaccine acceptance among college students in South Carolina and different factors that play a role in their vaccine uptake decision according to their levels of acceptance. Tailored vaccine promotion messages should address specific concerns among the refusal and hesitancy groups. School could play a positive role in vaccine campaign since the reluctancy group considered that recommendation by their school was important in their decision making. Health educators also need to pay particular attention to the refusal group who do not value duration of protection or authoritative advice as much as their counterparts in their vaccine decision making.

## Introduction

The novel coronavirus disease 2019 (COVID-19) pandemic has become a public health crisis worldwide and led massive impacts on health, economics, and individuals’ life (McKinsey & Company, 2020; Wang et al., 2020). The United States (US) has been experiencing the largest burdens of the pandemic. As of December 3, 2020, there have been a total number of 273,924 fatalities due to COVID-19 in the US, which is at the highest ranking in the world (Johns Hopkins University and Medicine, 2020a). To better control the pandemic, growing attention has been paid to COVID-19 vaccination. Since early 2020, global researchers have made efforts in developing and testing vaccines against COVID-19. It is estimated that a safe and effective COVID-19 vaccine will be available in late 2020 or early 2021 (Lurie, Saville, Hatchett, & Halton, 2020). Despite the availability, the success of COVID-19 vaccination would strongly depend on individuals’ vaccine acceptance. To achieve population immunity and significantly halt the spread of COVID-19, it is suggested a critical (minimum) herd-immunity threshold of 67% among general population (Kwok, Lai, Wei, Wong, & Tang, 2020). However, a recent global investigation on COVID-19 vaccine acceptance suggested a challenge to reach this threshold, showing that nearly 30% of participants would refuse or hesitate to take a COVID-19 vaccine when it is available (Lazarus et al., 2020). In order to develop effective COVID-19 vaccine promotion strategies, it is important to understand what factors would contribute to decision making of COVID-19 vaccination and whether these factors would differ between individuals who intend to take the vaccine and those who do not.

Vaccination decisions can be influenced by multiple factors. Vaccination theoretical frameworks, such as Increasing Vaccination Model, posit that vaccine uptake could be determined by factors from three aspects, including individual cognitions, social processes, and practice issues (Brewer, Chapman, Rothman, Leask, & Kempe, 2017; MacDonald, 2015). Factors of individual cognitions include individuals’ beliefs or attitudes towards vaccination, such as perceived efficacy or benefits of vaccines, safety concerns (e.g., side effects), and perceptions on characteristics of vaccines (e.g., ways to take vaccine and countries in which vaccines are made). Factors of social processes refer to interpersonal interactions on attitudes and perceptions of vaccination. An example of such factors is recommendations from significant others, such as family members and health authorities. Practice issues focus on factors that directly affect vaccination behaviors, such as vaccine availability, accessibility, and vaccination cost.

Several COVID-19 vaccination studies have reported associations between some of these factors (i.e., beliefs and attitudes, safety concerns, and provider recommendations) and COVID-19 vaccine acceptance (Fisher et al., 2020; Gadoth et al., 2020; Kasting, Head, Hartsock, Sturm, & Zimet, 2020; Reiter, Pennell, & Katz, 2020). However, most studies were conducted among healthcare providers or general population and limited literature have examined these factors in other groups at high risk of COVID-19. Also, it has not been well studied whether these factors could be weighed differently in vaccine decision making by individuals with different levels of COVID-19 vaccine acceptance. Vaccine literature has highlighted the value to investigate the patterns of weighing factors in vaccination decision making in different groups because individuals who hesitate or refuse to take vaccines may show different vaccine belief systems (Smith, 2017). Such knowledge can inform tailored vaccine promotion interventions or vaccine communication campaigns for people with different vaccine acceptance levels (acceptance, hesitancy, and refusal) to achieve a successful COVID-19 vaccination.

Young adults (aged 18 to 30 years) should be engaged in the campaigns of COVID-19 vaccination. Although older adults have been prioritized for COVID-19 prevention and treatment because of the elevated risk for severe illness with COVID-19 among this group, existing evidence has shown a comparable risk of COVID-19 infection in young adults. For example, data from the US, Geneva, and Switzerland consistently showed that seroprevalence of SAR-CoV-2 antibodies in young adults (9.9% to 10.9%) were similar in older adults (e.g., 55 to 64 years: 7.4% to 11.9%) (Guilamo-Ramos, Benzekri, Thimm-Kaiser, Hidalgo, & Perlman, 2020; New York State Governor’s Office, 2020; Stringhini et al., 2020). In addition, young adults are at high risk for COVID-19 transmission given that they are less likely to comply with preventive practices, including hand washing and social distancing, compared to other age groups (Czeisler et al., 2020; Zhang et al., 2020).

Among young adults, college students in the Southern US could be exposed to a higher risk of COVID-19 for two reasons. First, the COVID-19 epidemic is prevalent and serious in the South. As of October 29, 2020, the positivity of COVID-19 was 5.9% in South Carolina, and such a rate is higher than the threshold (5%) suggested by WHO for reopening (Johns Hopkins University and Medicine, 2020b). Second, a great number of college students in the South have been back to schools since majority of colleges have been reopened in Fall 2020. Many students attend to in-person classes and are living with roommates in apartments near campuses. In addition to COVID-19 vulnerability, it is important to note that the Southern States historically had a relatively lower vaccination rate for other infectious diseases than other states. For example, an investigation from eight universities in North Carolina revealed that only 14% to 30% of college students reported receiving influenza vaccines (Poehling, Blocker, Ip, Peters, & Wolfson, 2012), and such rates were significantly lower than the goal of 50% vaccine coverage suggested by the American College Health Association (ACHA) (ACHA, 2016). Taken together, college students in the South should be targeted for COVID-19 vaccination promotion. An investigation on their COVID-19 vaccine acceptance and the factors influencing their vaccination decision making could be critical to inform strategies for promoting COVID-19 vaccine uptakes among this population.

Given that limited research has addressed COVID-19 vaccine acceptance among college students in the South and factors influencing their vaccination decision-making, the current study aimed to (1) explore the proportions of college students who would accept, hesitate, or refuse to take a COVID-19 vaccine (i.e., level of vaccine acceptance); and (2) examine whether the factors that may affect vaccine acceptance were weighed differently in vaccination decision making by college students according to their levels of COVID-19 vaccine acceptance.

## Methods

### Participants and procedure

Online survey data were collected among a convenience sample of college students in South Carolina between September 2020 and October 2020. Data were collected using RedCap, a web-based survey platform which has been widely used in public health studies (Paris & Hynes, 2019). College students were invited to the study through student listservs in various colleges and departments on campus. Participants were eligible for the study if they were: (1) 18 years of age or older; and (2) full-time students currently enrolled in a university in South Carolina.

Potential participants could access the online survey via the hyperlink provided in the invitation email. An online informed consent was presented to the participants before they began the survey. The online consent covered necessary study information such as study purposes, confidentiality protection, voluntary nature of participation, and survey procedure. The survey typically took 20 minutes to complete. Upon completion, participants were offered an opportunity to win a $25 Amazon e-gift card through a prize draw. A total of 1370 college students responded to the survey. Ten e-gift cards were given away through a random drawing. Data from 308 participants were excluded due to the low completion rate (less than 50% of the survey), resulting in a total sample size of 1062 in the current study. The research protocol was approved by the Institutional Review Board at University of South Carolina.

### Measures Demographics

Participants were asked to provide their demographic information including gender (0 = female, 1 = male), age (years), annual family income (from < $10,000 to ≥ $100,000), race/ethnicity (e.g., White/Caucasian, Black/Africa American), and school year (i.e., Freshman, Sophomore, Junior, Senior, first or second year in their master or doctoral program). Due to a low proportion of sample in certain categories, we dichotomized race/ethnicity (0 = White/Caucasian, 1 = non-White/Caucasian) and school year (0 = undergraduate, 1 = graduate) for the purpose of data analysis in the current study.

### COVID-19 vaccine acceptance

Participants answered one question asking their likelihood of taking a COVID-19 vaccine when the vaccine is available (i.e., “How likely will you take a COVID-19 vaccine when it is available”). The question was rated using a five-point Likert scale (1 = definitely not take it, 2= not likely to take it, 3 = I don’t know, 4 = likely to take it, and 5 = definitely take it). Based on their responses, participants were divided into three groups including (1) refusal group, (participants with answers of ‘1’ or ‘2’); (2) hesitancy group (participants with answers of ‘3’); and (3) acceptance group (participants with answers of ‘4’ or ‘5’).

### Factors associated with COVID-19 vaccination decision making

A self-developed measure was used to assess factors that might influence the decision making of the COVID-19 vaccine uptake. This measure consisted of 12 items asking participants the extent to which certain COVID-19 vaccine-related factors would be important in their COVID-19 vaccination decision making. Items included efficacy of the COVID-19 vaccines, the duration of COVID-19 vaccine protection, negative consequences of COVID-19 vaccines (e.g., “side effects”), COVID-19 vaccine characteristics (e.g., “ways the vaccines will be administered [oral or injection]”), access to COVID-19 vaccines (e.g., “local availability of the vaccines”), and authoritative advice (e.g., “whether it is recommended by my doctors”). Participants rated items on a 7-point scale ranging from 1 (the least important) to 7 (the most important). Cronbach’s alpha of this measure was 0.75 in the current study.

### Statistical analysis

Data screening was conducted in terms of proper coding, univariate outliers (z scores), and normality (skewness and kurtosis). Descriptive statistics were reported for demographic variables (i.e., age, gender, race/ethnicity, annual family income, and school year) and COVID-19 vaccine acceptance. Univariate analyses, including analysis of variance (ANOVA) (for continuous variables) and chi-square test (for categorical variables), were utilized to examine the difference of demographic variables by COVID-19 vaccine acceptance levels (i.e., refusal, hesitancy, and acceptance).

Multivariate analysis of covariance (MANCOVA) was employed to examine the differences on the perceived importance of 12 COVID-19 vaccine-related factors by the levels of COVID-19 vaccine acceptance. Demographic variables were included as covariates in the analysis. Bartlett’s Test of Sphericity was used to determine if dependent variables (perceived importance of COVID-19 vaccine-related factors) were sufficiently correlated to where the correlation matrix diverges significantly from the identity matrix. To examine the homogeneity of variance assumption for MANCOVA, the Box’M test was employed. As suggested by Meyers, Gamst, and Guarino (2016), the coefficient of Omnibus effect was determined by the results of Box’M test. Wilk’s lambda was used when Box’M test suggested a multivariate support for the homogeneity of variance assumption (p > .05), while Philai’s trace was used when results did not support the homogeneity assumption.

Several follow-up analysis of covariance (ANCOVA) tests were employed on perceived importance of COVID-19 vaccine-related factors if MANCOVA suggested significant differences. Post-hoc Fisher’s Least Significant Difference (LSD) tests were then employed only on the factors that showed significant group differences by ANCOVA results. All statistical analyses were performed using SPSS software version 26.

## Results

### Descriptive statistics and univariate analyses

As shown in Table 1, participants were mostly female (79.8%) and White/Caucasian (85.9%). More than 50% participants were undergraduates (12.2% Freshmen, 10.5% Sophomore, 12.4% Junior, and 17.1% Senior). Nearly 40% participants reported annual family income of $100,000 or more.

**Table 1.** Demographic analyses between COVID-19 vaccination acceptance groups (*n* = 1062)

|  |  | Overall | Refusal group | Hesitancy group | Acceptance group | Group comparisons |  |
| --- | --- | --- | --- | --- | --- | --- | --- |
|  |  |  |  |  |  | *F/Chi-square* | *p-value* |
| *n* |  | 1062 | 258 (24.3%) | 160 (15.1%) | 644 (60.6%) |  |  |
| Demographics |  |  |  |  |  |  |  |
| Age, *Mean (SD)* |  |  | 24.64 (7.53) | 23.21 (6.03) | 23.64 (6.41) | 2.77 | 0.063 |
| Gender |  |  |  |  |  |  |  |
|  | Female | 848 (79.8%) | 214 (83.3%) | 134 (83.8%) | 500 (77.9%) | 4.93 | 0.085 |
|  | Male | 211 (19.9%) | 43 (16.7%) | 26 (16.3%) | 142 (22.1%) |  |  |
| Race/Ethnicity<sup>a</sup> |  |  |  |  |  |  |  |
|  | White/Caucasian | 912 (85.9%) | 216 (83.7%) | 136 (85.0%) | 560 (87.0%) | 1.71 | 0.425 |
|  | Black/Africa American | 71 (6.7%) |  |  |  |  |  |
|  | Hispanic/Latino | 32 (3.0%) |  |  |  |  |  |
|  | Asian | 85 (8.0%) |  |  |  |  |  |
|  | American Indian/Alaskan Native | 6 (0.6%) | 42 (16.3%) | 24 (15.0%) | 84 (13.0%) |  |  |
|  | Native Hawaiian/ other Pacific Islander | 3 (0.3%) |  |  |  |  |  |
|  | Other | 6 (0.6%) |  |  |  |  |  |
| Annual family income |  |  |  |  |  |  |  |
| | < \$10,000 | 63 (5.9%) | 15 (5.8%) | 8 (5.1%) | 40 (6.3%) | 13.23 | 0.104 |
| | \$10,000 to \$24,999 | 102 (9.6%) | 15 (5.8%) | 18 (11.5%) | 69 (10.8%) | | |
| | \$25,000 to \$49,999 | 161 (15.2%) | 47 (18.3%) | 29 (18.6%) | 85 (13.4%) | | |
| | \$50,000 to \$100,000 | 306 (28.8%) | 84 (32.7%) | 46 (29.5%) | 176 (27.7%) | | |
| | >\$100,000 | 417 (39.3%) | 96 (37.4%) | 55 (35.3%) | 266 (41.8%) | | |
| School year<sup>b</sup> |  |  |  |  |  |  |  |
|  | Freshman | 130 (12.2%) |  |  |  | 3.51 | 0.173 |
|  | Sophomore | 112 (10.5%) |  |  |  |  |  |
|  | Junior | 132 (12.4%) | 143 (55.6%) | 90 (57.0%) | 323 (50.3%) |  |  |
|  | Senior | 182 (17.1%) |  |  |  |  |  |
|  | Masters student, first year | 100 (9.4%) |  |  |  |  |  |
|  | Masters student, second year or above | 107 (10.1%) | 114 (44.4%) | 68 (43.0%) | 319 (49.7%) |  |  |
|  | Doctoral student, first year | 78 (7.3%) |  |  |  |  |  |
|  | Doctoral student, second year or above | 216 (20.3%) |  |  |  |  |  |

<sup>a</sup> Race/Ethnicity was dichotomized into 0 (White/Caucasian) and 1 (non-White/Caucasian) for analyses<sup>b</sup> School year was dichotomized into 0 (undergraduate) and 1 (graduate) for analyses.

In terms of COVID-19 vaccine acceptance, 60.6% participants were considered as acceptance group (i.e., ‘definitely’ or ‘likely to take COVID-19 vaccines’), 24.3% were considered as refusal group (i.e., ‘definitely not’ or ‘not likely to take COVID-19 vaccines’), and 15.1% were considered as hesitancy group (i.e., ‘I don’t know’). Univariate analyses suggested none of the demographic variables significantly differed across three groups.

### Multivariate analyses

Results of MANCOVA were presented in Table 2. Bartlett’s Test of Sphericity was significant □^2^= 3104.84, *p* < .001), suggesting that the correlation matrix among dependent variables was sufficiently diverged from the identity matrix. The Box’M test was significant (Box-M = 380.42, *p* < .001), failing to provide multivariate support for the homogeneity of variance. As such, Pillai’s trace was used to determine the omnibus effects. After accounting for demographics, MANCOVA revealed a statistically significant omnibus effect for COVID-19 vaccine acceptance (Pillai’s trace = 0.27, *F* [24, 1926] = 12.54, *p* < .001). Therefore, follow-up ANCOVAs were run to identify which of the 12 factors would differ across COVID-19 vaccine acceptance groups in terms of perceived importance.

**Table 2.**
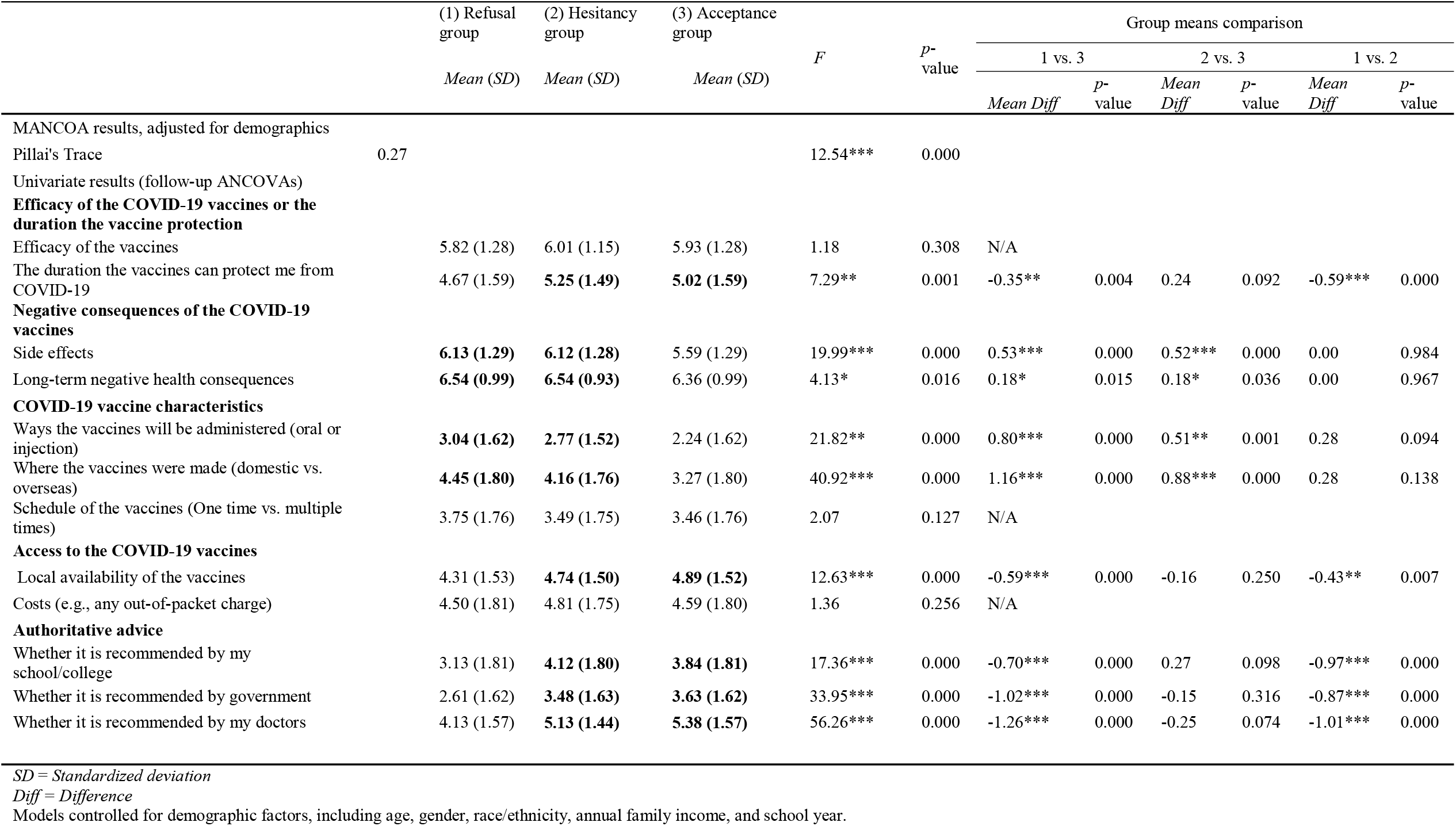
Factors associated with COVID-19 vaccine acceptance among college students (*n* = 1062)

Follow-up ANCOVAs suggested significant group differences on nine of the 12 factors in terms of perceived importance, including ‘side effects’ (*F* [2, 973] = 19.99, *p* < .001), ‘long-term negative health consequences’ (*F* [2, 973] = 4.13, *p* = .02), ‘ways the vaccines will be administered’ (*F* [2, 973] = 21.82, *p* < .001), ‘where the vaccines are made’ (*F* [2, 973] = 40.92, *p* = .001), ‘local availability of the vaccines’ (*F* [2, 973] = 12.63, *p* < .001), ‘whether it is recommended by my school/college’ (*F* [2, 973] = 17.36, *p* < .001), ‘whether it is recommended by government’ (*F* [2, 973] = 33.95, *p* < .001), ‘whether it is recommended by my doctors’ (*F* [2, 973] = 56.26, *p* < .001), and ‘the duration the vaccines can protect me from COVID-19’ (*F* [2, 973] = 7.29, *p* = .001).

LSD tests were performed for these nine factors to identify the pair-wise group differences according to their levels of COVID-19 vaccine acceptance (Table 2). Results suggested different patterns across three groups in terms of perceived importance of each factor. Compared to the refusal group, the acceptance group reported higher scores on ‘local availability of the vaccines’ (*p* = .007), ‘whether it is recommended by the school/college’ (*p* < .001), ‘whether it is recommended by government’ (*p* < .001), ‘whether it is recommended by my doctors’ (*p* < .001), and ‘the duration the vaccines can protect me from COVID-19’ (*p* < .001). Relative to the acceptance group, the refusal group had greater scores on ‘side effects’ (*p* < .001), ‘long-term negative health consequences’ (*p* = .015), ‘ways the vaccines will be administered’ (*p* < .001), and ‘where the vaccines will be made’ (*p* < .001).

The hesitancy group reported relatively higher scores in all domains of factors. Compared to the refusal group, the hesitancy group had greater scores on factors in authoritative advice (‘whether it is recommended by my school/college’, ‘government’, or ‘doctors’) (*p*s < .001), access to the COVID-19 vaccines (‘local availability of the vaccines’), and ‘the duration the vaccines can protect me from COVID-19’ (*p* < .001). Compared to the acceptance group, the hesitancy group had higher scores on factors in negative consequences of the COVID-19 vaccines (‘side effects’ and ‘long-term negative health consequences’) (*p*s < .05) and COVID-19 vaccine characteristics (‘ways the vaccine will be administered’ and ‘where the vaccines are made’) (*p*s < .01). Although with a marginally statistical significance, the hesitancy group reported a higher score on ‘whether it is recommended by my school/college’) (*p* = .098) and ‘the duration the vaccines can protect me from COVID-19’) (*p* = .092) than the acceptance group.

## Discussion

The current study examined the COVID-19 vaccine acceptance and explored how a number of COVID-19 vaccine-related factors could be weighed differently in their vaccination decision making according to the levels of vaccine acceptance among college students in South Carolina. To the best of our knowledge, the current study was one of the first attempts to document acceptance rate of COVID-19 vaccines and factors influencing future COVID-19 vaccine uptake among the college students in the Southern US.

Our results suggested that 60.6% of college students were likely or definitely to take a COVID-19 vaccine when available. This acceptance rate was lower than 71.5% in a global sample and 75.4% in a US sample (Lazarus et al., 2020). This finding was aligned with previous studies on influenza vaccination that reported a significantly lower flu shot coverage in young adults than other age groups from 2010 to 2019 (Centers for Disease Control and Prevention, 2020b). The low acceptance rate may reflect optimistic bias in young adults. Compared with other age groups, young adults were more likely to underestimate severity of the disease and perceive low susceptibility of COVID-19 infection (Pasion, Paiva, Fernandes, & Barbosa, 2020; Wise, Zbozinek, Michelini, Hagan, & Mobbs, 2020). The vaccine refusal and hesitancy in college students could be a critical concern given the need of a minimum immunity level of 67% to achieve population immunity (Kwok et al., 2020). Hence, developing interventions to improve COVID-19 vaccine acceptance and promote vaccine uptake among college students in the South merits a high attention.

Our findings suggested that perceived importance of COVID-19 vaccine-related factors in vaccination decision making differed by COVID-19 vaccine acceptance levels among college students. College students who would hesitate or refuse to take a COVID-19 vaccine should be targeted for vaccine promotion interventions. Compared with other groups, hesitancy group considered authoritative advice from health authorities (health providers and government) important in their vaccine decision making. This finding is consistent with a US national survey study suggesting that a provider recommendation could boost COVID-19 vaccine intentions (Head, Kasting, Sturm, Hartsock, & Zimet, 2020). Given the strong influence of authoritative advice, CDC has released a guidance regarding effective COVID-19 vaccine conversations in clinics and highlighted the influences of healthcare providers on recommending patients to take COVID-19 vaccines (“give your strong recommendation”) (Centers for Disease Control and Prevention, 2020a). It is also worth noting that, although only a slight difference, hesitancy group weighed more on the recommendation by schools than acceptance group. Such a finding implies that schools can play an important role in promoting COVID-19 vaccine acceptance among college students. A campus-wide COVID-19 vaccine communication is warranted by emphasizing college’s perspective on COVID-19 vaccination and providing recommendations from health authorities. In addition, hesitancy group had similar concerns as acceptance group in terms of the duration of COVID-19 vaccine protection (i.e., “the duration of the vaccines can protect me from COVID-19”) and access to the COVID-19 vaccines (e.g., “local availability of the vaccines”).

These findings imply that vaccination decisions would be influenced by the beliefs about protection functionality of COVID-19 vaccine and the convenience of taking a vaccine when it is available. COVID-19 vaccine campaign on college campuses should be tailored to address these two issues by providing clear information regarding the duration of vaccine protection and making the COVID-19 vaccine accessible to the college students.

Our findings suggested that hesitancy and refusal groups viewed negative consequences of COVID-19 vaccines (‘side effects’ and ‘long-term consequences’) and COVID-19 vaccine characteristics (‘ways the vaccines will be administered’ and ‘where the vaccines are made’) as important factors in deciding whether they will take COVID-19 vaccine. Concerns on safety and quality of COVID-19 vaccines could be barriers against vaccine acceptance. Extant literature suggests that safety concerns and mistrust in vaccines contribute to vaccine hesitancy, and these concerns may become salient when the vaccine is new and rapidly developed (Dror et al., 2020; Karafillakis et al., 2016). Vaccine literature has suggested that misinformation regarding vaccines and a lack of sophisticated knowledge of immunization may induce anxiety and perceived uncertainty, leading to an overestimation of the potential side effects of the vaccines (Bliss & Morrison, 2020; Dubé, Gagnon, Nickels, Jeram, & Schuster, 2014; Karafillakis & Larson, 2017; Larson, 2018). To reduce college students’ concerns on safety and quality of COVID-19 vaccines, evidence-based health communication in colleges should address misinformation about vaccines (e.g., ‘fact check’) and deliver vaccine knowledge using population-appropriate languages.

It is also important to note that refusal group reported high scores on negative consequences or characteristics of COVID-19 vaccines but low scores on the duration of vaccine protection and authoritative advice. This finding implies that college students with low vaccine acceptance outweighed potential negative consequence of vaccination than the benefits of vaccination. To provoke their attentions to other aspects of COVID-19 vaccination, message-framing techniques would be warranted for college-wide health communications. For example, loss-framed messages that emphasize the consequences of not taking vaccines have been found to be effective in increasing willingness of getting vaccinated among young adults (Lee & Cho, 2017). In addition, recent literature has also highlighted the value of prosocial-framed messages, which could enhance individuals’ attentions to prosocial benefits of COVID-19 vaccination such as the protection of communities and significant others (Chou & Budenz, 2020; Jordan, Yoeli, & Rand, 2020).

There are several methodological limitations in the current study. First, data were collected from a convenience sample of college students in South Carolina. Findings in the current study may not be generalized to students in other states. Second, self-report data may be subject to response bias, such as social desirability. Third, cross-sectional data cannot draw causal inferences. Fourth, measures on factors associated with COVID-19 vaccination decision making were self-developed and have not been validated. Future research should use a random sample, apply a longitudinal design, and validate self-developed measures.

Despite these limitations, the current study is one of the first attempts to explore COVID-19 vaccine acceptance among college students in the South. We identified an acceptance rate of 60.6%, which is lower than that in general population and merits a public health attention since young adults have comparable risk of COVID-19 with other age groups. Our findings show factors associated with vaccination decision making were weighed differently by college students with different vaccine acceptance levels, which may have important implications to public health practices. Acceptance-enhancing interventions or vaccine communications in colleges could benefit from tailoring contents to the patterns of decision making. College leaders and healthcare providers need to be aware of their important role in promoting COVID-19 vaccination. The success of vaccination may strongly rely on young adults’ participations. Policy makers, healthcare practitioners, colleges in the South need to work together and make efforts in enhancing COVID-19 vaccine acceptance among college students.

## Data Availability

Data availability can be contacted through the corresponding author.

## Notes

### Competing Interest Statement

The authors have declared no competing interest.

### Funding Statement

Research reported in this publication was supported by the National Institutes of Health under Award Number of NIH R01MH0112376-3S1 and R01AI127203-4S1. The content is solely the responsibility of the authors and does not necessarily represent the official views of the National Institutes of Health.

### Author Declarations

The study protocol was approved by the Institute Review Board at University of South Carolina.

